# Neuro-immune, metabolic, and oxidative pathways in depression due to hypothyroidism and Hashimoto’s thyroiditis

**DOI:** 10.1101/2025.07.25.25332222

**Authors:** Sahira Qasim Al-Baldawi, Hussein Kadhem Al-Hakeim, Ikram Khémiri, Michael Maes

**Author notes:** **Corresponding author:** Prof. Dr. Michael Maes, M.D., Ph.D. Sichuan Provincial Center for Mental Health Sichuan Provincial People’s Hospital, School of Medicine, University of Electronic Science and Technology of China, Chengdu 610072, China. https://scholar.google.co.th/citations?user=1wzMZ7UAAAAJ&hl=th&oi=ao.

## Abstract

**Background:** Hypothyroidism is linked to depression and several metabolic alterations, including insulin resistance, dyslipidemia, and oxidative stress. This study investigates the impact of hormones, autoimmunity, metabolic, and antioxidant indicators on the severity of depression in patients with hypothyroidism.

**Methods:** Forty-six patients with hypothyroidism and seventy-four with Hashimoto’s thyroiditis participated in this study, along with sixty healthy controls. Patients were categorized based on the Hamilton Depression Rating Scale (≥ 17) into those with depression and those without. The enzyme-linked immunosorbent assay method was employed to evaluate blood insulin and selenoprotein P (SePP). Graphite furnace atomic absorption spectrophotometry was employed to quantify serum selenium concentrations. Serum zinc and lipid profile indicators were measured using spectrophotometry.

**Results:** Hypothyroidism and Hashimoto’s thyroiditis are linked to increased atherogenicity, insulin resistance, and reduced antioxidant defenses, including selenium, SePP, and zinc. Both cohorts with thyroid dysfunctions demonstrate slight elevations in depressive symptoms. Individuals with hypothyroidism and heightened depressive symptoms demonstrated augmented insulin resistance, raised atherogenic indices, and markedly reduced levels of SePP relative to those with milder depressive symptoms. Elevated levels of thyroid-stimulating hormone and atherogenic index of plasma best predicted the severity of depression in hypothyroid patients.

**Conclusions:** The findings indicate that depression due to hypothyroidism is largely influenced by abnormalities in thyroid hormones, thyroid-stimulating hormone, metabolic pathways, and diminished antioxidant defenses. The observed results may be explained by the established impact of these hormones and biomarkers on cerebral functions, resulting in major depressive disorder.

## Introduction

Depression is one of the most prevalent mental problems associated with thyroid disease (Peng et al., 2023). The incidence of depression in hypothyroid individuals differs, with certain studies indicating rates reaching 80% in particular communities (Ahmed et al., 2024). In a study, almost 25% of those diagnosed with depression demonstrated irregular thyroid function (Kafle et al., 2020).

Hypothyroidism is an established risk factor for depression, with processes related to the impact of free thyroxine (FT4), free triiodothyronine (FT3), and thyroid-stimulating hormone (TSH) on mood regulation (Nuguru et al., 2022, Bode et al., 2021). Enhancing thyroid function in hypothyroid individuals may mitigate affective symptoms such as anxiety and depression (Fanaei et al., 2022, Demartini et al., 2014). Thyroid hormone receptors are located in many brain regions, including the amygdala, hippocampus, and cerebral cortex, all of which are associated with the development of mental diseases (Williams, 2008). Reduced levels of FT3 and FT4 affect cerebral blood flow and may lead to neurocognitive impairments and neurodegeneration. Hypothyroidism may hinder axodendritic growth, synaptogenesis, and neuronal migration in the cerebral cortex and hippocampus, resulting in morphological abnormalities, including fewer granular cells and mossy fibers in the hippocampus and reduced hippocampal volume (Bernal et al., 2024, Cooke et al., 2014, Li et al., 2024). In individuals with Hashimoto’s disease, the intensity of depression has a substantial correlation with the concentrations of anti-thyroid peroxidase antibodies (TPOAb) and anti-thyroglobulin antibodies (TgAb) (Karagun, 2024, Aydemir, 2023). Animal studies indicate that Hashimoto’s thyroiditis can provoke neuroinflammation and affective changes, implying that TgAb and TPOAb antibodies may initiate inflammatory responses in the brain, resulting in mood disorders (Cai et al., 2018).

In addition to the influence of thyroid hormones on depression, various additional factors may contribute to comorbid depression, including aberrations in metabolic processes, trace elements, and antioxidant levels. Reduced concentrations of thyroid hormones in hypothyroidism correlate with heightened insulin resistance (IR) (Abdel-Gayoum, 2016, Unnikrishnan et al., 2023), partially attributable to the involvement of thyroid hormones in the modulation of insulin receptors and glucose metabolism. The insufficiency of thyroid hormones in Hashimoto’s thyroiditis results in increased serum concentrations of total cholesterol (TC), low-density lipoprotein cholesterol (LDLc), and triglycerides (TG) (Liu and Peng, 2022, Matshar, 2024). Dyslipidemia is prevalent in hypothyroid patients, with studies indicating its occurrence in up to 90% of individuals (Mansfield et al., 2022, Harrar et al., 2024). Triglycerides and the atherogenic index of plasma (AIP) were markedly elevated, whereas high-density lipoprotein cholesterol (HDLc) was considerably diminished in hypothyroid disease (Pekgör et al., 2025).

Blood concentrations of critical trace elements, particularly selenium and zinc, may directly influence thyroid function in individuals with Hashimoto’s thyroiditis and overt hypothyroidism (Rasic- Milutinovic et al., 2017, Rostami et al., 2024, Lu et al., 2023). Selenium is essential for the production of selenoproteins, including glutathione peroxidase and iodothyronine deiodinases, which are integral to thyroid hormone synthesis, metabolism and antioxidant defense within the thyroid gland (Wang et al., 2023, Dudinskaya, 2024, Köhrle and Gärtner, 2009). Furthermore, the thyroid gland depends on selenoproteins to mitigate oxidative stress by neutralizing reactive oxygen species, therefore maintaining the structural and functional integrity of thyrocytes (Pesic et al., 2015, Wang et al., 2023). Selenium may affect mood regulation and has been linked to depressed symptoms (Santos et al., 2022). Selenium is predominantly conveyed in plasma by selenium-containing protein P (SePP1), which delivers selenium to peripheral organs, particularly the thyroid gland. Decreased SePP levels correlate with increased depression symptomatology (Birģele et al., 2025). Variations in SePP levels may contribute to the pathophysiology of depression and anxiety through pathways linked to oxidative stress and inflammation (Barchielli et al., 2022). Zinc deficiencies can disrupt thyroid function, leading to reduced thyroid hormone levels and altered enzyme activities, which may exacerbate iodine deficiency and contribute to thyroid dysfunction (Khanam, 2018, Senyushkina and Troshina, 2021). Zinc is crucial for the optimal operation of TPO, the activity of deiodinases, and the structural integrity and functionality of the T3 receptor (Severo et al., 2019, Nóbrega, 2019). Lowered zinc is also a marker of major depressive disorder and treatment-resistant depression (Maes et al., 1997).

The aforementioned indicates that neuro-immune processes (autoimmune mechanisms potentially impacting the brain), metabolic issues (insulin resistance and atherogenicity), and oxidative stress (reduced zinc, selenium, and selenium-binding protein) are implicated in the pathogenesis of hypothyroid disease and Hashimoto’s disease. Recent reviews indicate that intertwined aberrations in neuro-immune, metabolic, and oxidative stress (NIMETOX) pathways are pivotal in the etiology and pathophysiology of depression (Maes et al., 2025a).

However, it remains uncertain whether autoimmune factors (TPOAb and TgAb), metabolic conditions (insulin resistance and atherogenicity), lower antioxidant defenses (selenium, selenium-binding protein, zinc), or hormonal alterations (FT3, FT4, TSH) are the primary determinants of depressive symptoms in hypothyroid disorder or Hashimoto’s disease. Hence, this study seeks to investigate the link between depression and selenium, SePP1, zinc, atherogenic indices, and insulin resistance in patients with hypothyroidism. The specific hypothesis is that these indicators are partially responsible for the manifestation of depressive symptoms due to hypothyroid disease.

## Subjects and Methods

### Participants

The current study included forty-six clinical cases of hypothyroid disease and seventy-four patients diagnosed with Hashimoto’s thyroiditis. The subjects were recruited at Al-Sadr Medical City, Najaf Governorate, Iraq, between September 2024 and November 2024. The study was conducted in accordance with Iraqi and international ethical and privacy norms. The Medical Ethics Committee at the University of Kufa approved this research ethically, Reference #: MEC-96, by the International Guidelines for Human Research Protection as outlined by the Declaration of Helsinki. Informed permission was obtained from each participant prior to their inclusion in the study.

The diagnosis of hypothyroid disease was made following the 10th version of the International Classification of Diseases and Related Health Problems, Clinical Modification 2024 (ICD-10-CM, Diagnosis Code E03.9). Patients exhibiting hypothyroid disease and Hashimoto’s thyroiditis in the euthyroid or subclinical stage were excluded. Sixty apparently healthy individuals (29 females and 31 males) were assigned to a control group. Their ages and sex ratio were matched to those of the patients.

The patient’s assessment encompassed a thorough medical history to identify any systemic illnesses that could affect the evaluated parameters. The current investigation excluded any subjects with significant systemic diseases, particularly cardiovascular or renal diseases, as well as manifest inflammatory conditions, diabetes mellitus type 1, viral hepatitis, infectious disease, renal illness, cardiovascular disease, cancer, hypertension, autoimmune disorders such as inflammatory bowel disease, rheumatoid arthritis, multiple sclerosis, psoriasis, and neurodegenerative disease such as Parkinson’s and Alzheimer’s disease. Furthermore, all subjects with lifetime psychiatric disorders such as depression, schizophrenia, bipolar disorder, autism, substance abuse disorders (except tobacco use disorder) posttraumatic stress disorder were excluded from participating. Pregnant and lactating women were excluded.

The Hamilton Depression Rating Scale (HAMD), 17 symptom version, was utilized to assess the severity of depression. Sixty depressed patients (HT+Depression) exhibiting moderate to severe depression (HAMD>17) were compared to sixty patients with lower HAMD scores.

### Assays

Five milliliters of fasting blood samples were aspirated from each participant using disposable needles and plastic syringes. The blood was allowed to coagulate for ten to fifteen minutes at room temperature. After that, the blood was centrifuged at 1200 Xg for five minutes to separate the serum. Serum was subsequently transferred to new, disposable Eppendorf tubes and stored at -35°C in a blood bank deep freezer for the duration of the experiment. The enzyme-linked immunosorbent assay (ELISA) approach was used to assess blood insulin and SePP using ELISA kits bought from Nanjing Pars Biochem Co., Ltd. (Nanjing, China). All ELISA kits have intra-assay coefficients of variation (CV) ≤ 10%. The electrochemiluminescence immunoassay was employed on Elecsys and Cobas e immunoassay analyzers for the quantitative measurement of FT3, FT4, TSH, TPOAb, and TgAb in serum samples. The Elecsys FT4 II assay (ROCHE-eLabDoc, Germany) employs an antigen-specific antibody conjugated with a Tris(2,2’-bipyridyl)ruthenium (II) complex (Ru(bpy)) for antigen detection. The graphite furnace atomic absorption spectrophotometry experiments utilized the Shimadzu AA-6300 device from Japan to estimate serum selenium levels. Serum zinc level was quantified spectrophotometrically using kits provided by Giesse Diagnostics (Rome, Italy). Total cholesterol (TC), triglycerides (TG), fasting blood glucose (FBG), and HDL cholesterol were quantified with kits provided by Spinreact^®^, Gerona, Spain. Low-density lipoprotein cholesterol (LDLc) was calculated using Friedewald’s formula, as outlined below: LDLc = TC - (TG/2.19 + HDLc) (Friedewald et al., 1972). Castelli’s Risk Index-1 (CRI-1) was derived from TC/HDL, whereas Castelli’s Risk Index-2 (CRI-2) was derived from LDLc/HDLc (Castelli et al., 1983). The AIP is determined by the logarithmic transformation of the molar ratio of triglycerides to high-density lipoprotein cholesterol (Log(TG/HDLc)) (Dobiásová et al., 2011). The Homeostasis Model Assessment 2 (HOMA2) calculator^©^, developed by the Diabetes Trials Unit at the University of Oxford (https://www.dtu.ox.ac.uk/homacalculator/download.php), was utilized to compute β-cell function (HOMA%B), insulin sensitivity (HOMA%S), and insulin resistance (HOMA2-IR) based on fasting blood insulin and glucose concentrations.

### Biostatistical analysis

A one-way ANOVA was employed to compare scale variables among groups. Protected Fisher’s Least Significant Difference (LSD) was employed for pairwise comparisons following ANOVA for scale variables. Contingency tables (χ^2^tests) were employed to evaluate the relationships between categorical variables. Pearson’s product-moment correlation was employed to examine the associations among scale factors. We performed a binary logistic regression analysis using the diagnosis of hypothyroid disease (HT) or HT + HAMD>17 (HT+MDD) as the dependent variable, compared to controls or patients without major depression (HT-MDD), with biomarkers serving as explanatory variables. The odds ratio with 95% confidence intervals was calculated, along with the predictive accuracy, sensitivity, and specificity. Multiple regression analysis was employed to identify the key biomarkers predicting the HAMD score through manual and stepwise automatic methods (p-to-entry of 0.05 and p-to-remove of 0.06). We calculated the standardized beta coefficients for each significant explanatory variable utilizing t statistics with precise p-values, alongside the model F statistics and the total variance explained (R²), which served as an effect size. Additionally, the analysis was evaluated for homoscedasticity through the White and Breusch-Pagan tests, and for collinearity issues using VIF and tolerance metrics. All statistical analyses were conducted using SPSS 30 (IBM-USA).

## Results

### Demographic, clinical, and biomarker data

The results in **Table 1** show no significant difference among the two hypothyroid samples and control study groups in age, BMI, sex, smoking, marital status, and HDLc. Both hypothyroid patient groups have a significant increase in TSH, HAMD, FBG, FBI, TC, LDLc, and the three atherogenic indices as compared with controls. Both patient groups show a significant decrease in FT3, FT4, selenium, SePP, and zinc in comparison with the controls. Hashimoto’s thyroiditis group has a significant increase in TPOAb and TgAb compared with the two other study groups. HDL was lower in Hashimoto’s thyroiditis compared with controls. All patients with hypothyroid disorder were treated with L-thyroxine, 39 patients with propranolol, and 21 patients (all with Hashimoto’s thyroiditis with glucocorticoids). Using GLM analysis we were unable to find any effects of the use of propranolol and glucocorticoids on the results of this study.

**Table 1.** Demographic and biochemical features of patients with hypothyroidism, Hashimoto’s thyroiditis, and healthy controls (HC)

| Variables | HC (n=60) | Hypothyroidism (n=46) | Hashimoto's thyroiditis (n=74) | F/$\chi^2$ | p |
| --- | --- | --- | --- | --- | --- |
| Age Years | 40.0±10.3 | 41.6±9.87 | 41.03±9.043 | 0.40 | 0.670 |
| Sex Female/Male | 29/31 | 18/28 | 37/37 | 1.45 | 0.485 |
| Smoking No/Yes | 42/18 | 28/18 | 55/19 | 2.43 | 0.296 |
| Single/Married | 11/49 | 13/33 | 20/54 | 1.84 | 0.398 |
| Body mass index kg/m<sup>2</sup> | 28.24±3.61 | 28.29±3.04 | 27.95±3.17 | 0.20 | 0.820 |
| HAMD score | 5.18±1.68<sup>B,C</sup> | 13.07±6.89<sup>A</sup> | 13.23±6.62<sup>A</sup> | 41.01 | <0.001 |
| TSH uIU/mL | 2.72±0.73<sup>B,C</sup> | 7.61±2.30<sup>A</sup> | 8.11±2.51<sup>A</sup> | 132.17 | <0.001 |
| FT3 pM | 4.26±0.51<sup>B,C</sup> | 3.44±1.13<sup>A</sup> | 3.45±1.10<sup>A</sup> | 14.57 | <0.001 |
| FT4 pM | 17.10±2.81<sup>B,C</sup> | 10.10±1.41<sup>A</sup> | 9.84±1.41<sup>A</sup> | 258.84 | <0.001 |
| TPOAb IU/ml | 18.0±8.8<sup>C</sup> | 17.5±9.1<sup>C</sup> | 410.0±183.7<sup>A,B</sup> | 239.94 | <0.001 |
| TGAb IU/ml | 66.4±28.5<sup>C</sup> | 70.7±30.9<sup>C</sup> | 304.0±325.9<sup>A,B</sup> | 27.34 | <0.001 |
| Selenium ng/ml | 136.2±30.8<sup>B,C</sup> | 83.7±22.9<sup>A</sup> | 75.7±25.2<sup>A</sup> | 93.97 | <0.001 |
| Selenoprotein pg/mL | 201.7 ±65.6<sup>B,C</sup> | 150.7 ±56.9<sup>A</sup> | 142.9 ±67.4<sup>A</sup> | 15.26 | <0.001 |
| Zinc mg/L | 0.85±0.13<sup>B,C</sup> | 0.70±0.14<sup>A</sup> | 0.70±0.15<sup>A</sup> | 22.31 | <0.001 |
| Fasting blood glucose mM | 5.20±0.48<sup>B,C</sup> | 5.77±0.47<sup>A</sup> | 5.81±0.52<sup>A</sup> | 29.31 | <0.001 |
| Fasting blood insulin pM | 54.51±12.10<sup>B,C</sup> | 77.17±27.69<sup>A,C</sup> | 65.29±23.54<sup>A,B</sup> | 14.20 | <0.001 |
| HOMA2-IR index | 1.03±0.22<sup>B,C</sup> | 1.48±0.52<sup>A,C</sup> | 1.26±0.45<sup>A,B</sup> | 16.04 | <0.001 |
| Total cholesterol (TC) mM | 4.81±0.35<sup>B,C</sup> | 5.32±0.55<sup>A</sup> | 5.43±0.53<sup>A</sup> | 29.42 | <0.001 |
| High-density lipoprotein (HDL) mM | 0.99±0.11<sup>C</sup> | 0.96±0.13 | 0.94±0.13<sup>A</sup> | 2.36 | 0.097 |
| Low-density lipoprotein (LDL) mM | 3.38±0.37<sup>B,C</sup> | 3.73±0.53<sup>A</sup> | 3.83±0.50<sup>A</sup> | 16.03 | <0.001 |
| Castelli risk index 1 (TC/HDL) | 4.92±0.60<sup>B,C</sup> | 5.64±0.87<sup>A</sup> | 5.85±0.82<sup>A</sup> | 25.39 | <0.001 |
| Castelli risk index 2 (LDL/HDL) | 3.47±0.58<sup>B,C</sup> | 3.97±0.80<sup>A</sup> | 4.13±0.73<sup>A</sup> | 15.45 | <0.001 |
| Atherogenic index of plasma | -0.005±0.077<sup>B,C</sup> | 0.160±0.102<sup>A</sup> | 0.182±0.111<sup>A</sup> | 65.92 | <0.001 |

All results are shown as mean±SD. $\chi^2$ : results of analysis of contingency analysis (df=2); F: results of analysis of variance (all: df=2/177). <sup>A,B,C</sup>: protected post-hoc analysis. HOMA2IR: Homeostatic Model Assessment 2 for Insulin Resistance, TSH: thyroid stimulating hormone, FT3: Free triiodothyronine, FT4: free thyroxine, TPOAb: thyroid peroxidase autoantibodies, and TgAb: thyroglobulin autoantibodies.

**Table 2** shows the measurements of the same variables now in controls and hypothyroid patients divided according to severity of depression (cutoff value: HAMD ≥ 17). The HAMD score was significantly different among the three groups. SePP was significantly lower in the HT+MDD than in the control and HT-MDD groups. FBI, FBG, TC, and the three atherogenic indices were significantly different between the three study groups and increased from controls to HT-MDD to HT+MDD. HDL was significantly lower in HT+MDD than in the two other study groups.

**Table 2.** Demographic and biochemical features of hypothyroid patients with (HT+MDD) and without depression (HT-MDD), and healthy controls (HC)

| Variables | HC<br>n=60 | HT-MDD<br>n=60 | HT+MDD<br>n=60 | F/$\chi^2$ | p |
| --- | --- | --- | --- | --- | --- |
| Age years | 40.0±10.3 | 41.0±10.5 | 41.6±8.1 | 0.40 | 0.671 |
| Sex Female/Male | 29/31 | 28/32 | 27/33 | 0.13 | 0.935 |
| Body mass index kg/m<sup>2</sup> | 28.24±3.61 | 28.13±3.22 | 28.04±3.02 | 0.06 | 0.942 |
| HAMD score | 5.18±1.68<sup>B,C</sup> | 6.92±1.27<sup>A,C</sup> | 19.42±3.07<sup>A,B</sup> | 784.73 | <0.001 |
| TSH IU/mL | 2.72±0.73<sup>B,C</sup> | 7.69±2.07<sup>A</sup> | 8.15±2.74<sup>A</sup> | 131.98 | <0.001 |
| FT3 pM | 4.26±0.51<sup>B,C</sup> | 3.34±0.95<sup>A</sup> | 3.55±1.24<sup>A</sup> | 15.37 | <0.001 |
| FT4 pM | 17.10±2.81<sup>B,C</sup> | 10.13±1.35<sup>A</sup> | 9.75±1.46<sup>A</sup> | 260.12 | <0.001 |
| TPOAb IU/ml | 18.0±8.7<sup>B,C</sup> | 235.4±220.7<sup>A</sup> | 283.7±256.9<sup>A</sup> | 31.41 | <0.001 |
| TGAb IU/ml | 66.4±28.5<sup>B,C</sup> | 211.3±255.3<sup>A</sup> | 217.9±305.1<sup>A</sup> | 8.30 | <0.001 |
| Selenium ng/ml | 136.2±30.8<sup>B,C</sup> | 81.5±26.4<sup>A</sup> | 76.0±22.4<sup>A</sup> | 92.64 | <0.001 |
| Selenoprotein pg/mL | 201.7 ±65.6<sup>C</sup> | 155.5 ±59.9<sup>C</sup> | 136.3 ±65.9<sup>A,B</sup> | 16.59 | <0.001 |
| Zinc mg/L | 0.85±0.13<sup>B,C</sup> | 0.71±0.16<sup>A</sup> | 0.69±0.13<sup>A</sup> | 22.93 | <0.001 |
| Fasting blood glucose mM | 5.20±0.48<sup>B,C</sup> | 5.62±0.43<sup>A,C</sup> | 5.97±0.51<sup>A,B</sup> | 39.64 | <0.001 |
| Fasting blood insulin pM | 54.51±12.10<sup>B,C</sup> | 65.51±22.76<sup>A,C</sup> | 74.18±27.96<sup>A,B</sup> | 12.10 | <0.001 |
| HOMA2-IR index | 1.03±0.22<sup>B,C</sup> | 1.26±0.43<sup>A,C</sup> | 1.44±0.53<sup>A,B</sup> | 14.64 | <0.001 |
| Total cholesterol (TC) mM | 4.81±0.35<sup>B,C</sup> | 5.30±0.51<sup>A,C</sup> | 5.48±0.55<sup>A,B</sup> | 31.28 | <0.001 |
| High-density lipoprotein (HDLc) mM | 0.99±0.11<sup>C</sup> | 0.99±0.15<sup>C</sup> | 0.90±0.08<sup>A,B</sup> | 11.51 | <0.001 |
| Low-density lipoprotein (LDLc) mM | 3.38±0.37<sup>B,C</sup> | 3.75±0.49<sup>A</sup> | 3.83±0.54<sup>A</sup> | 15.85 | <0.001 |
| Castelli risk index 1 (TC/HDLc) | 4.92±0.60<sup>B,C</sup> | 5.42±0.80<sup>A,C</sup> | 6.12±0.73<sup>A,B</sup> | 42.01 | <0.001 |
| Castelli risk index 2 (LDLc/HDLc) | 3.47±0.58<sup>B,C</sup> | 3.85±0.77<sup>A,C</sup> | 4.28±0.70<sup>A,B</sup> | 21.41 | <0.001 |
| Atherogenic index of plasma | -0.005±0.077<sup>B,C</sup> | 0.089±0.076<sup>A,C</sup> | 0.267±0.061<sup>A,B</sup> | 211.08 | <0.001 |

All results are shown as mean±SD. F: results of analysis of variance (all: df=2/177). $\chi^2$ : results of analysis of contingency analysis (df=2); <sup>A,B,C</sup>: protected post-hoc analysis. HOMA2IR: Homeostatic Model Assessment 2 for Insulin Resistance, TSH: thyroid stimulating hormone, FT3: Free triiodothyronine, FT4: free thyroxine, TPOAb: thyroid peroxidase autoantibodies, and TgAb: thyroglobulin autoantibodies.

### Results of binary logistic regression analysis

To delineate the best predictors of HT versus controls, we have performed binary logistic regression analyses with HT as the dependent variable (controls as reference group) and the biomarkers as explanatory variables (**Table 3**). The first regression shows that the increase in FBG, AIP, TC, coupled with a decrease in selenium, substantially discriminated between the two study groups (χ^2^=176.100, df=4, p<0.001). The Nagelkerke effect size was 0.867, and the classification precision was 94.4%, with a sensitivity of 93.3% and a specificity of 95.0%.

**Table 3.** Results of binary logistic regression analyses with hypothyroid disease (HT) or HT with depression (HT+MDD) as dependent variables and biomarkers as explanatory variables.

| Dichotomies | Explanatory variables | $\beta$ | SE | Wald | df | p | OR | 95% CI |
| --- | --- | --- | --- | --- | --- | --- | --- | --- |
| 1. HT vs. controls | Selenium | -0.064 | 0.015 | 17.504 | 1 | <0.001 | 0.938 | 0.910-0.967 |
|  | Fasting blood glucose | 2.226 | 0.830 | 7.189 | 1 | 0.007 | 9.261 | 1.820-47.132 |
|  | Atherogenic index of plasma | 2.295 | 0.665 | 11.898 | 1 | <0.001 | 9.926 | 2.694-36.573 |
|  | Total cholesterol | 1.958 | 0.637 | 9.435 | 1 | 0.002 | 7.082 | 2.031-24.696 |
| 2. HT+MDD vs. HT-MDD | Atherogenic index of plasma | 3.829 | 0.656 | 34.074 | 1 | <0.001 | 46.010 | 12.721-166.412 |

HT-MDD: hypothyroid disorder without MDD

The second regression analysis examined the discrimination between HT+MDD versus HT- MDD and found that HT+MDD was best predicted by increased AIP levels (χ^2^=98 117, df=1, p<0.001) with an effect size of 0.745 (Nagelkerke R^2^), and overall accuracy of 90.8%, with a sensitivity of 90.0%, and specificity of 91.7%.

### Results of multivariate regression analysis

**Table 4** presents the results from multiple regression analyses with the HAMD score, AIP, TSH and FT4 as dependent variables and the other biomarkers as explanatory variables. Regression #1 indicates that a substantial portion of the variation in the HAMD score (59.1%) was accounted for by the regression on AIP and TSH. **Figure 1** illustrates the partial regression of the HAMD total score on the AIP. Regression #2 indicates that a substantial portion of the variation in AIP levels (42.6%) was elucidated by the regression on FBG, TPOAb, and HOMA2IR (all positively correlated), as well as FT4 and selenium (all negatively correlated). Regression #3 indicates that 43.2% of the variation in TSH levels is accounted for by hyperglycemia (positively correlated) and selenium (negatively correlated). As much as 47.8% of the variation in FT4 levels was elucidated by selenium and zinc levels (both positively correlated) and FBG levels (negatively correlated). **Figure 2** illustrates the partial regression of FT4 on selenium levels.

**Figure 1.**
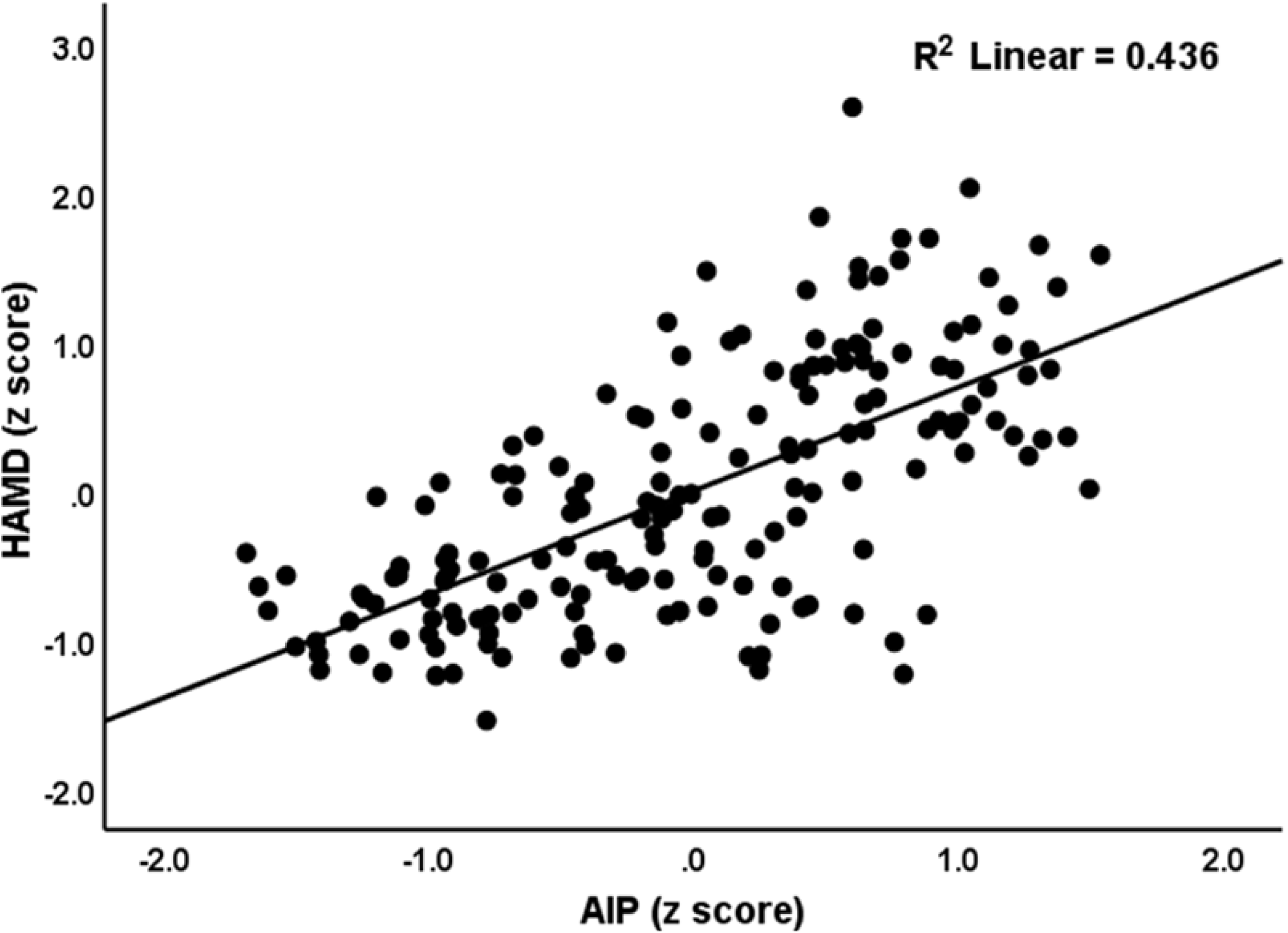
Partial regression of the Hamilton Depression Rating Scale (HAMD) score on the atherogenic index of plasma (AIP) (p<0.001).

**Figure 2.**
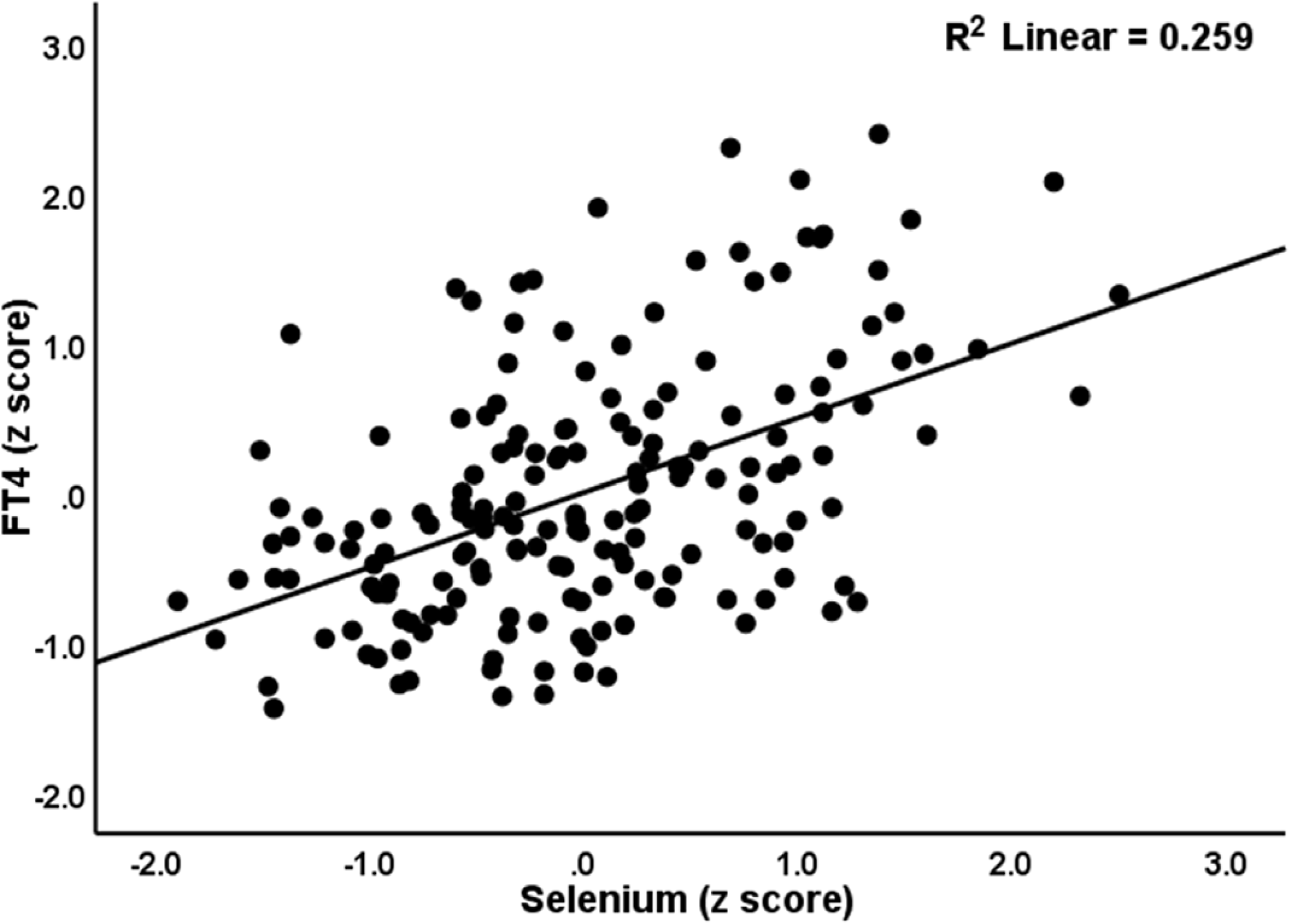
Partial regression of free thyroxine (FT4) on serum selenium levels (p<0.001).

**Table 4.** Results of multiple regression with the total Hamilton Depression Rating Scale (HAMD) score, atherogenic index of plasma (AIP), thyroid secreting hormone (TSH), and free thyroxine (FT4) levels as dependent variables, and serum biomarkers as explanatory variables.

| Dependent Variables | Explanatory Variables | $\beta$ | t | p | F model | df | p | R<sup>2</sup> |
| --- | --- | --- | --- | --- | --- | --- | --- | --- |
| #1. HAM-D | **Model** |  |  |  | 128.06 | 2/177 | <0.001 | 0.591 |
|  | Atherogenic index of plasma | 0.692 | 12.31 | <0.001 |  |  |  |  |
|  | Thyroid stimulating hormone | 0.132 | 2.34 | 0.020 |  |  |  |  |
| #2. AIP | **Model** |  |  |  | 25.87 | 5/174 | <0.001 | 0.426 |
|  | Free thyroxine | -0.216 | -2.61 | 0.010 |  |  |  |  |
|  | Fasting blood glucose | 0.218 | 3.29 | 0.001 |  |  |  |  |
|  | Selenium | -0.164 | -2.16 | 0.032 |  |  |  |  |
|  | TPOAb | 0.174 | 2.61 | 0.010 |  |  |  |  |
|  | HOMA2-IR | 0.155 | 2.51 | 0.013 |  |  |  |  |
| #3. TSH | **Model** |  |  |  | 67.22 | 2/177 | <0.001 | 0.432 |
|  | Selenium | -0.471 | -7.70 | <0.001 |  |  |  |  |
|  | Fasting blood glucose | 0.313 | 5.11 | <0.001 |  |  |  |  |
| #4. FT4 | **Model** |  |  |  | 53.82 | 3/176 | <0.001 | 0.478 |
|  | Selenium | 0.499 | 7.95 | <0.001 |  |  |  |  |
|  | Fasting blood glucose | -0.231 | -3.89 | <0.001 |  |  |  |  |
|  | Zinc | 0.134 | 2.23 | 0.027 |  |  |  |  |

HOMA2IR: Homeostatic Model Assessment 2 for Insulin Resistance, TPOAb: thyroid peroxidase autoantibodies.

## Discussion

### NIMETOX pathways in hypothyroid disease

This study’s initial conclusion indicates that hypothyroid illness and Hashimoto’s thyroiditis are associated with heightened atherogenicity, insulin resistance, and diminished antioxidant defenses, including selenium, SePP, and zinc. A significant portion of the variance in the AIP index was elucidated by the combined influences of IR, diminished selenium and FT4 levels, and heightened thyroid autoimmunity. Elevated basal TSH and diminished FT4 levels correlated with serum selenium and fasting blood glucose, but reduced zinc was further linked to decreased FT4. The data indicates that metabolic disorders and diminished antioxidant defenses are integral components of hypothyroid illness. Decreased T3 and T4 levels in hypothyroidism can disrupt lipid metabolism, resulting in elevated atherogenic lipoproteins, including LDL and triglycerides, and diminished HDL levels (Ekinci, 2025). These modifications are evident in elevated atherogenicity indices, including AIP (Tao et al., 2024). Thyroid hormones are crucial for regulating glucose metabolism and influencing the expression of genes related to gluconeogenesis, glycogen synthesis, and lipid metabolism, which can lead to compromised insulin signaling and glucose uptake in tissues (Teixeira et al., 2020). Selenium deficiency can hinder the function of selenoenzymes critical for thyroid hormone synthesis and antioxidant protection, resulting in heightened oxidative stress and diminished thyroid hormone production, thereby contributing to thyroid inflammation and autoimmune thyroiditis (Dudinskaya, 2024, Zuo et al., 2021). SePP serves as the primary selenium transporter and functions as a potent antioxidant that neutralizes peroxynitrite (Steinbrenner et al., 2016). Zinc is crucial for the synthesis and metabolism of thyroid hormones, including T3 and T4 (Severo et al., 2019), and serves as a potent antioxidant (Lee, 2018).

### NIMETOX pathways in depression due to hypothyroid disease

Our research indicates that both groups with thyroid dysfunctions exhibit mild increases in depressive symptoms. Additionally, we discovered that hypothyroid individuals with moderately elevated depressive symptoms exhibited increased insulin resistance, atherogenicity indices, as well as significantly lower levels of antioxidants (SePP) in comparison to the HT group with milder depressive symptoms. In addition, we identified robust correlations between metabolic variables and diminished antioxidant defenses and disturbances in TSH and thyroid hormone levels. Consequently, our findings indicate that depression due to hypothyroidism is influenced by the interconnected disorders in TSH/thyroid hormone levels and NIMETOX biomarkers.

It is known that hypothyroid patients are at an increased risk of developing depression, as evidenced by prior research (Bode et al., 2021, Roa Dueñas et al., 2024, Demartini et al., 2014). Cross-sectional and longitudinal correlations between thyroid function and depression have been observed in longitudinal investigations. More severe depressive symptoms are associated with lower levels of FT4 and FT3 (Roa Dueñas et al., 2024, Nuguru et al., 2022, Ma et al., 2024, Nilkantham and Singh, 2024), and higher TSH levels (Hamed et al., 2021, Khan et al., 2022, Kim et al., 2015, Liu et al., 2022). Furthermore, Hashimoto’s thyroiditis is associated with substantial correlations between depression and TPOAb levels (Degner et al., 2014). Individuals with autoimmune thyroid disease and elevated TPOAb may be at an increased risk of developing anxiety and depression (Morina, 2024).

Insulin resistance is known to be a notable risk factor for depression, especially in non-obese persons (Rhee et al., 2023). Although insulin resistance is not increased in patients with MDD, it can contribute to depression severity, probably by increasing neurotoxicity (Maes et al., 2025b). In major depression, increased atherogenicity is a critical metabolic factor in depressive phenomenology. This is due to the increased neurotoxicity that results from elevated free cholesterol and reduced reverse cholesterol transport, which is accompanied by a decrease in HDL and specific HDL-bound antioxidant enzymes (Maes et al., 2025a, Maes et al., 2025b). The latter contributes to more profound deficiencies in total antioxidant defenses, which result in increased oxidative damage to lipids and proteins (Maes et al., 2025a, Maes et al., 2025b).

### Mechanistic explanations

Our results show that depression due to hypothyroidism is influenced by disorders in TSH/thyroid hormones and NIMETOX pathways, which may be explained by the known effects of these hormones and biomarkers on brain functions.

Decreased serum levels of FT3 and FT4 are associated with diminished cerebral blood flow perfusion in association with cognitive deficits (Hu et al., 2016). In fact, optimal thyroid hormone levels are essential for sustaining adequate cerebral circulation (Fani et al., 2022). Moreover, elevated anti-thyroid antibodies in the cerebrospinal fluid of individuals with depression reinforce the hypothesis that central autoimmunity may have a role in mood disorders (Dersch et al., 2020).

Immune-inflammatory pathways might play a key role as persistent low-grade inflammation linked to insulin resistance might result in neurodegeneration and intensify depressive symptoms by influencing brain energy metabolism and fostering insulin insensitivity (Leonard and Wegener, 2020). Insulin resistance may lead to heightened neurotoxicity and diminished neuroplasticity, ultimately compromising synaptic connections and brain circuitry, especially in areas essential for cognition and emotional regulation, such as the frontal and subcortical regions (Rosenblat et al., 2014, Maes et al., 2023). Moreover, insulin resistance is associated with mitochondrial dysfunction, marked by diminished oxidative activity and heightened oxidative stress, which further intensifies neuronal injury (Kleinridders et al., 2015).

Individuals with mood disorders exhibit markedly elevated atherogenic indices, particularly the AIP and the Castelli Risk Index 2 (CRI-2) (Jirakran et al., 2025). Increased atherogenicity affects neuronal homeostasis which may result in neuronal cell toxicity and death (Cheon, 2023). Indices of immune-linked neurotoxicity and atherogenicity were highly correlated with the clinical phenotypic characteristics of depression (Maes et al., 2024). Furthermore, oxidative stress, frequently intensified in atherogenic conditions, might aggravate neuronal injury, neuronal inflammation and neuronal death (Bhatt et al., 2020).

Selenium and SePP are associated with the pathophysiology of depression due to their effects on thyroid metabolism and antioxidant properties. Selenium insufficiency has been associated with both hypothyroid disorders and depression, as it is essential for the production of selenocysteine-containing proteins that govern thyroid hormone metabolism and antioxidant defense mechanisms (Sun et al., 2021, Sajjadi et al., 2022). In hypothyroid disorder, selenium metabolism and thyroid function interact in the etiology of depressive symptoms (Sun et al., 2021, Soheili-Nezhad et al., 2023). Interestingly, a meta-analysis showed that selenium supplementation might decrease TPOAb levels in individuals receiving T4 therapy (Wichman et al., 2016). Reduced serum SePP levels may serve as a possible biomarker for depression (Birģele et al., 2025). Our results suggest that reduced levels of selenium and SePP in hypothyroidism may exacerbate affective symptoms secondary to thyroid disorders. Lower SePP levels have been linked to oxidative stress and inflammation, perhaps contributing to the pathophysiology of affective symptoms (Barchielli et al., 2022).

Insufficient zinc levels facilitate the onset of depression via multiple interrelated biochemical pathways. Zinc deficiency may cause increased oxidative stress, compromised neurogenesis, and heightened neurodegeneration, which are all fundamental characteristics of depression (Szewczyk et al., 2011).

## Limitations

This study possesses some limitations that require acknowledgment. The case-control design prohibits any causal inferences on the observed changes in NIMETOX indicators linked to the onset of depression in hypothyroid populations. Secondly, despite our meticulous selection of pertinent biomarkers, the study did not encompass a broader spectrum of immunological (e.g., cytokine levels) and oxidative stress (e.g., lipid peroxidation) pathways that may be involved in hypothyroid-related depression. Third, given that the study was conducted in Iraq, it may lack generalizability to other nations or cultures. Fourth, while there were no effects of treatments with propranolol and glucocorticoids on the results of the present study, possible effects of L-thyroxine treatment could not be ruled out.

## Conclusion

In individuals with hypothyroid disease, the current study found increased insulin resistance, dyslipidemia and atherogenicity, and deficits in antioxidants including selenium, SePP, and zinc. Depression due to hypothyroid disease was associated with more pronounced aberrations in these biomarkers. In patients with hypothyroidism, the best predictors of depression are a rise in fasting blood glucose, atherogenicity, as well as a drop in selenium. As a result, metabolic disorders and lowered antioxidative defenses are additional drug targets to treat patients with depression due to hypothyroid disease.

## Author’s contributions

All the contributing authors have participated in the preparation of the manuscript.

## Data availability statement

The database created during this investigation will be provided by the corresponding author (MM) upon a reasonable request once the authors have thoroughly used the data set.

